# Preliminary evaluation of the safety and efficacy of oral human antimicrobial peptide LL-37 in the treatment of patients of COVID-19, a small-scale, single-arm, exploratory safety study

**DOI:** 10.1101/2020.05.11.20064584

**Authors:** Hanlin Zhang, Yiming Zhao, Xiaoxiao Jiang, Yuying Zhao, Li Yang, Li Chen, Meng Dong, Zhe Luan, Chunlong Yan, Jianwei Jiao, Chaoyue Zhao, Hongyue Li, Wei Chen, Cong Feng, Le Tian, Enqiang Qin, Jinsong Mu, Congyong Li, Tianshu Zeng, Shibo Feng, Shufang Wang, Xizhou Guan, Tanshi Li, Haotian Yu, Aihua-Zheng, Wanzhu Jin, Gang Sun

**Affiliations:** Key Laboratory of Animal Ecology and Conservation Biology, Institute of Zoology, Chinese Academy of Sciences, Beijing100101, China; University of Chinese Academy of Sciences, Beijing 100049, China; Department of Gastroenterology and Hepatology, Hainan Hospital of PLA General Hospital, Sanya 572013, China; Department of Cardiovascular Medicine, 980 Hospital of PLA Joint Logistics Support Forces, Shijiazhuang 050082, China; Department of Endocrinology, Hankou Hospital of Wuhan City, Wuhan 430012, China; Department of Gastroenterology and Hepatology, the first medical center, Chinese PLA General Hospital, Beijing 100853, China; Agricultural College, Yanbian University, Yanji, 133002, China; Savaid Medical School, University of the Chinese Academy of Sciences, Beijing, 100049, China; State Key Laboratory of Integrated Management of Pest Insects and Rodents, Institute of Zoology, Chinese Academy of Sciences, Beijing 100101, China; CAS Center for Excellence in Biotic Interactions, University of Chinese Academy of Sciences, Beijing 100049, China; Emergency department, the first medical center, Chinese PLA General Hospital, Beijing 100853, China; Department of health and logistics support, Hainan Hospital of PLA General Hospital, Sanya 572013, China; Department of Hepatology, the fifth medical center, Chinese PLA General Hospital, Beijing 100853, China; Intensive Care Unit, the fifth medical center, Chinese PLA General Hospital, Beijing 100853, China; Department of Geriatric Gastroenterology, the sixth medical center, Chinese PLA General Hospital, Beijing 100048, China; Department of Endocrinology, Union Hospital, Tongji Medical College, Huazhong University of Science and Technology, Wuhan 430022, China; Department of Orthopaedics, Hankou Hospital of Wuhan City, Wuhan 430012, China; Department of Respiratory Medicine, the first medical center, Chinese PLA General Hospital, Beijing 100853, China

**Author notes:** **These authors contributed equally to this work** (Hanlin Zhang, Yiming Zhao, Xiaoxiao Jiang, Yuying Zhao, Li Yang, Li Chen, Meng Dong, Zhe Luan). **Contact information for Correspondence:** Tianshu Zeng M.D., Department of Endocrinology, Union Hospital, Tongji Medical College, Huazhong University of Science and Technology, Wuhan 430022, China; Hubei provincial Clinical Research Center for Diabetes and Metabolic Disorders, Wuhan 430022, China; Shibo Feng M.D., Department of Orthopaedics, Hankou Hospital of Wuhan City, Wuhan 430012, China; Shufang Wang M.D., Department of Gastroenterology and Hepatology, the first medical center, Chinese PLA General Hospital, Beijing 100853, China; Xizhou Guan M.D., Department of Respiratory Medicine, the first medical center, Chinese PLA General Hospital, Beijing 100853, China; Tanshi Li M.D., Emergency department, the first medical center, Chinese PLA General Hospital, Beijing 100853, China; Haotian Yu M.D., Department of health and logistics support, Hainan Hospital of PLA General Hospital, Sanya 572013, China; Aihua-Zheng Ph.D., State Key Laboratory of Integrated Management of Pest Insects and Rodents, Institute of Zoology, Chinese Academy of Sciences, Beijing 100101, China; CAS Center for Excellence in Biotic Interactions, University of Chinese Academy of Sciences, Beijing 100049, China; Wanzhu Jin Ph.D., Key Laboratory of Animal Ecology and Conservation Biology, Institute of Zoology, Chinese Academy of Sciences, Beijing 100101, China; Savaid Medical School, University of the Chinese Academy of Sciences, Beijing, 100049, China; Gang Sun Ph.D. & M.D., Department of Gastroenterology and Hepatology, the first medical center, Chinese PLA General Hospital, Beijing 100853, China;.

**Keywords:** COVID-19, SARS-COV2, LL-37, Safety and Efficacy Clinical Study

## Abstract

**Background& Aims:** The Coronavirus Disease 2019 (COVID-19) has become a global epidemic and has caused a lasting and huge loss of life security, economic development and social stability in more than 180 countries around the world. Unfortunately, there is still no specific treatment for COVID-19 till now, therefore, at this point, all potential therapies need to be critically considered. LL-37 is one of the best-studied human antimicrobial peptide (AMPs) that has a broad-spectrum activity against bacteria and viruses. The use of living, genetically modified organisms (GMOs) is an effective approach for delivery of therapeutic proteins. The aim of this study was to determine the safety and efficacy of the Lactococcus lactis which has been genetically modified to produce the therapeutic human antimicrobial peptide LL-37 (herein after referred to cas001) in the patients of COVID-19.

**Methods:** Firstly we constructed genetically modified food-grade probiotic, Lactococcus lactis, with sequence of seven tandem repeats of mature human LL-37 under control of the nisin-inducible nisA promoter to produce the cas001. A total of 20 healthy SD rats, half male and half female (There were five male and five female in the control group, the same in treatment group) were used to observe the acute toxic reaction and death after daily administration of cas001 for three weeks, which helps to provide necessary reference basis for clinical dose selection, verificaition of toxic reaction and possible target organs. According to the estimated clinical dosage of 1 × 10^8^CFU /kg/day, considering the conversion of body surface area, the dose for rats should be multiplied by 6.17 to 6 × 10^8^ CFU/kg/day. We administrated 100 times higher dose at 6 × 10^10^ CFU/ /kg/day to rats. In order to investigate the pharmacokinetics of cas001, male SD rats (body weight 250-300g, 1 × 10^10^ /animal, n=3) were given oral administration of LL-37 bacteria powder. The concentration of LL-37 in the blood before and after gavage was detected by ELISA kit (Hycult biotechnology Cat# HK321).

Human clinical study was approved by Ethics committee of Chinese PLA General Hospital (S2020-074-04) and a total of 11 patients with mild symptoms were enrolled in Wuhan hankou hospital and Huoshenshan hospital. They were enrolled voluntarily and all patients signed informed consent. Among them, there were 5 males and 6 females, aged 55 ± 12 (36-70) years old, and the duration from onset to medication enrollment was 35 ± 19 (5-68) days. 6 patients were nucleic acid positive and 5 patients were nucleic acid negative when they were enrolled. All patients received the oral drug cas001 treatment according to requirement(1 × 10^9^ CFU/capsule, 3 capsules/time, three times a day for 3weeks), with an average follow-up time of 33 ± 15 days (see table 1 for the results).

**Findings:** Western blot analysis shows that reasonable amount of LL-37 were induced by different concentrations of nisin, which means we have successfully constructed cas001. In the pre-clinical safety evaluation test, after three weeks administration of cas001, no adverse effects were observed on the rat’s body weight, food and water intake, hematological or serum biochemical parameters. The results showed that the LD_50_ of cas001 was higher than that of the 100 times of the expected clinical dose of 6 × 10^10^ CFU/day. These results showed that cas001 could be safe in animal experiments. In addition, rat pharmacokinetics results showed that the serum concentration of LL-37 reached peak 2 hours after gavage of cas001 and returned to basal level 6 hours after gavage. During study period, the volunteers did not feel any discomfort while taking the cas001 capsules, and two hours after oral administration, the concentration of LL-37 were increased in healthy volunteers.

cas001 shows definite effect in the improvement of gastrointestinal symptoms and is possible to have effects in improving the systemic symptoms and respiratory symptoms and may play a role in the improvement of results of nucleic acid test and lung CT test. 11 patients enrolled showed good compliance, tolerance, subjective feeling and actively interacted with the doctors. None of the patients had any adverse reactions.

**Conclusions:** Based on above observations, we conclude here that as an oral anti-viral agent, cas001 displayed good safety profiles. It is very hard to reach conclusion of clinical outcomes related to the cas001, although changes of several symptoms indicate encouraging findings.

## Introduction

On March 11, 2020, the director-general of World Health Organization (WHO) declared the severe acute respiratory syndrome coronavirus 2 (SARS-CoV-2) situation as a pandemic^1^. By May 4, 2020, the number of cases of COVID-19 had increased exponentially. There have been more than 3 million reported cases, 240 thousand death and the number of affected countries was more than 200. Recent report has demonstrated that there is less than 80% sequence similarity between SARS-CoV-2 and SARS-CoV^2^ and recent studies also pointed to the important role of ACE2 in mediating entry of SARS-CoV-2^2, 3^.

Currently, there is no specific treatment for COVID-19 and therapeutic options in response to the COVID-19 outbreak are urgently needed^4, 5^ For above reasons, there are several options can be considered for drug discovery: the first one is to screen anti-viral drugs that are approved by government agency with good safety profiles. The second one is to keep a close watch on virus entry and replication in the target cells. From this point of view, clinical trials are under way to test the safety and efficacy of recombinant human ACE2 in the treatment of COVID-19^6^. As early as February 2020, a Chinese research team discovered that remdesivir and chloroquine could effectively inhibit the SARS-CoV-2 in vitro by screening of the antiviral efficiency of five FDA-approved drugs^7^. In line with this regard, currently, at least 80 trials related to the chloroquine, hydroxychloroquine have been registered worldwide^8^. On the other hand, nearly twenty clinical trials regarding the remdesivir were also registered worldwide. In addition, by compassionate-use of remdesivir, clinical improvement was observed^9^. While we still need to accelerate and fine-tuning drug development to control infection rate, improve cure rate and decrease mortality of COVID-19.

LL- 37 is against a variety of viruses^10^, including the human acquired immunodeficiency virus (HIV-1)^11^, influenza a virus (IAV)^12^, respiratory syncytial virus (RSV)^13^, rhinovirus^14^, herpes simplex virus (HSV)^15^Zika virus^10^ and hepatitis c virus (HCV)^16^, etc., mainly through the interaction with cell membrane and virus shell to play antiviral effects. When the antimicrobial form binds to the cell membrane, the antimicrobial form disrupts the lipid layer, increasing the permeability of the cell membrane and leading to the outflow of material from the cell, thus killing the microorganism^17^. Antibacterial peptide LL-37 can induce antigen-presenting cells to produce a strong immune response, by producing and releasing type I interferon and promoting the maturation of dendritic cells, enhancing the transmission of viral dsDNA to toll-like receptors located near the nucleus, thus achieving an effective antiviral effect. There is a clinical study demonstrating that treatment with LL-37 for chronic leg ulcers was safe and well tolerated with the marked effect on healing predictors^18^.

Lactococcus lactis is non-pathogenic, non-colonizing bacterium and is one of the most commonly used microorganisms in the dairy industry and has an excellent safety record. Genetically modified probiotic Lactococcus lactis can serve as vectors for local delivery of biologically active molecules^19^. Several clinical studies were conducted with Lactococcus lactis, either bacteria itself on chronic rhino sinusitis (NCT04048174) and pre-hypertensive subjects (NCT02670811) or genetically modified lactococcus lactis that has been engineered to secrete human Interleukin-10 (hIL-10) which is now in a phase I trial to treat Crohn’s disease^20^, a Phase II study in Ulcerative Colitis (NCT00729872) or used as malaria vaccine hybrid GMZ2 (NCT00424944);What’s more, Lactococcus lactis secreting human TFF1 (Trefoil Factor 1) is in a phase II, double-blind, placebo-controlled, 2-arm, multi-center trial (NCT03234465). These studies all indicated that lactococcus lactis displays an excellent safety profiles.

The purpose of this study was to determine the safety and efficacy of the cas001 in the treatment of patients with COVID-19.

### Materials and animals

The food grade bacterial strains Lactococcus lactis cremoris, NZ3900 (Cat# VS-ELS03900-01) and plasmids pNZ8149 (Cat# VS-ELV00300-01) & pNZ8148(Cat# VS-ELV00200-01) were purchased from MoBiTec GmbH, Germany. In addition, fermenting materials were supported by Beijing Aoboxing Bio-Tech Co., LTD. Human LL-37 peptides came from GenScript Biotech Corporation (Cat# RP13323). Other biomedical reagents came from the Sigma-Aldrich company.

7-week-old male C57BL/6J mice and SD Rats were purchased from Vital River Laboratory Animal Technology. Co. LTD. Each mouse (n=5) was treated by 1.0E9 frozen-dried powder of chloromycetin resistant recombinant bacteria once and feces were collected for counting viable bacteria by pour plate method at 3, 9, 15, 27, 39 and 51 hr after administration respectively. After a week of adjustable feeding, male (n=10) and female (n=10) SD Rats were randomly separated into equal-number-individual groups respectively according to body weight and treated with 6.0E10/kg·day frozen-dried powder by gavage in the animal safety assessment. In the following two weeks, body weight, water drink, food intake, blood routine examination, serum biochemical analysis and daily observation were recorded to assess the risk of drug. For pharmacokinetics tracking, rats of same age were treated with same dose of recombinant bacteria (n=3), blood samples were collected from tail tip at 0, 1, 2, 4, 6 hr after gavage. Serum LL-37 concentration were detected by Hycult biotechnology human ELISA KIT (Cat# HK321).

### Ethics

Animals were housed under a 12/12 hr light/dark cycle with free access to food and water. Before sacrifice, animals were given an anesthetic and cervical dislocation rapidly to relieve pain. All animal studies were approved by the Institutional Animal Care and Use Committee of Institute of Zoology, Chinese Academy of Sciences.

The clinical human study was approved by the Ethics committee of Chinese PLA General Hospital (S2020-074-04) and all participants signed an informed consent according to the requirement of the Ethics committee. Based on safety concerns, the ethics committee requested a small exploratory clinical trial in which participants are patients with mild symptoms of COVID-19, and the study mainly focused on assessing the clinical safety prior to large-scale clinical study. At the same time, this study was registered on the west China website. (http://www.chictr.org.cn, ChiCTR2000030939).

### Plasmid cloning and transformation

The construction of DNA element is shown in the Figure 1B, DNA sequence was submitted to GenScript Biotech Corporation for gene synthesis and sub-cloning into pNZ8149. The pNZ8149-7×LL-37 recombinant vector were transferred into home-made NZ3900 competent cell through Gene PulserXcell™ electroporation system (BioRad, California, USA) to produce cas001.

**Figure 1.**
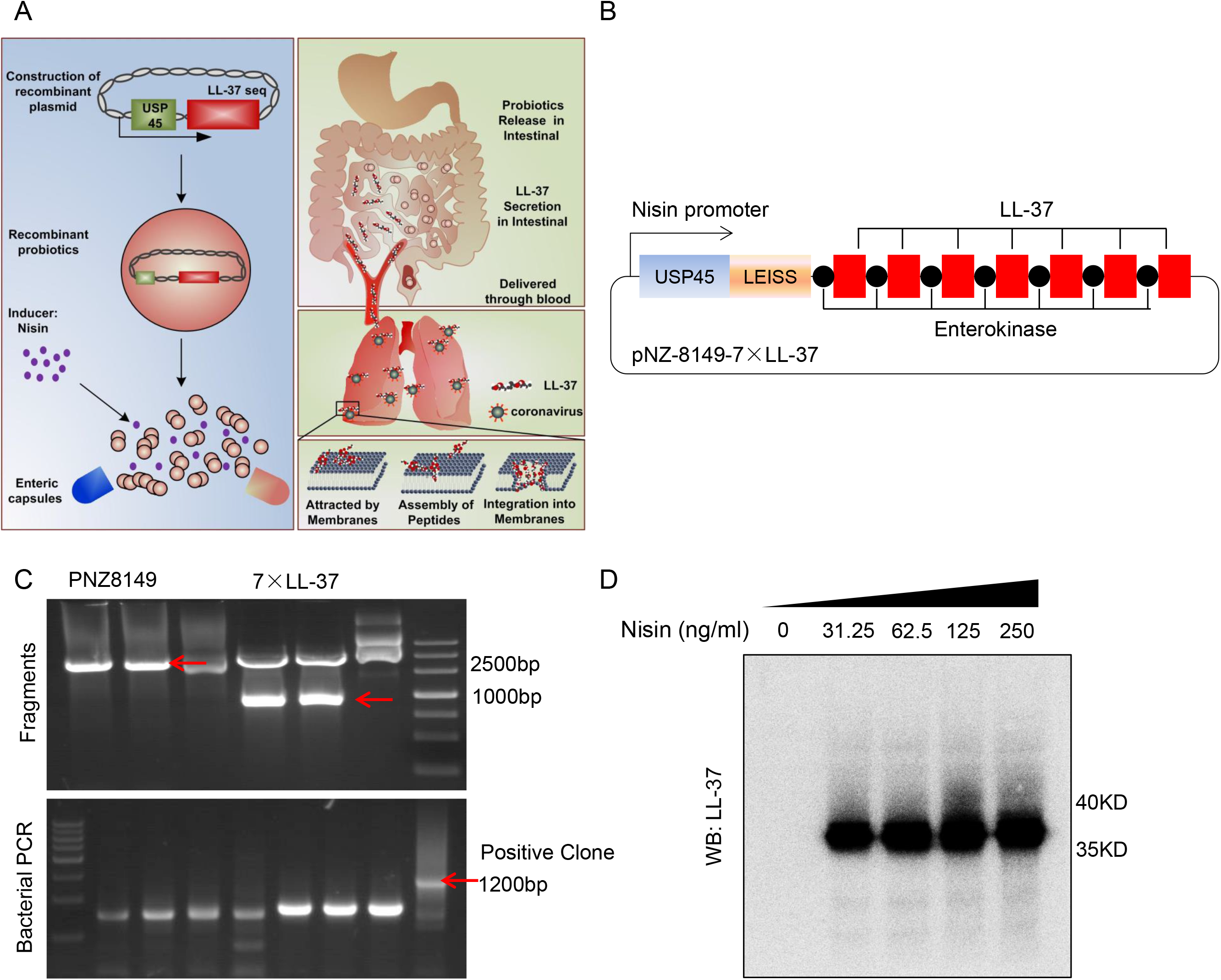
Design principles of oral antiviral drugs and recombinant vector construction. (A) Principle of oral administration of living biological drugs. (B) Amino acid sequence of recombinant 7×LL-37. (C) Agarose gel electrophoresis results of PCR fragments and suspected positive clone. (D) Western blotting detection of induced gene expression of recombinant vector in vitro.

### Cell culture

The cas001 were cultured at 30 °C up to 9-10 hours in mixed M17 media (0.5% Soya peptone, 0.25% peptone, 0.25% Casein peptone, 0.25% Yeast Extract, 0.5% Beef Extract, 0.05% Ascorbic acid, 1.9% β-Disodium Glycerophosphate and 0.025% MgSO47·H2O, adjusted to pH 7.2, replenishing 0.5% Lactose after being autoclaved before use) until the end of logarithmic growth phase. Before vacuum freeze-drying, these probiotics were collected by centrifuging and mixed with cryoprotectant (3.5% skim milk powder, 19.5% maltodextrin) in the right proportions for oral administration. Vero cell line, the courtesy of Aihua-Zheng, Ph.D, were cultured in DMEM high medium with 8%FBS, at 37°C and 5% CO2.

### Gene expression and western blotting

The cas001 were taken out from ultra-low temperature refrigerator for overnight activation twice at 30°C in M17 medium. Then the pure culture above was transferred into 10 ml fresh M17 medium for stationary culture until OD600 was around 0.4-0.6, different concentrations of nisin was added to induce the LL-37 fusion protein expression. Finally proteins in supernatant were concentrated by acidulated organic solvent precipitation for western blotting detection of target protein. The specific human LL-37 primary antibody came from Santa Cruz Biotech, CA, USA (Cat# sc-166770).

### Anti-virus infection experiment in vitro

GFP labeled-recombinant vesicular stomatitis virus-SARS (rVSV-SARS) and vesicular stomatitis virus-SARS-CoV2 (rVSV-SARS-CoV2) were gifts from Aihua-Zheng, Ph.D. The surface G-attachment glycoprotein of rVSV was specifically replaced by the spike protein of SARS or SARS-CoV2. The rVSV-SARS or rVSV-SARS-CoV2 were diluted into 2000 ffu/ml as working solution, then different doses of LL-37 were added into working solution respectively and incubated for 30 min at room temperature. Then vero cells copped in 96 well plate were changed the medium with this working solution and continued to culture for 2 hr at 37°C, each group was performed with three biological replicates. Afterwards vero cells were replaced with fresh medium including 20 mM NH4Cl to terminate virus infection for 14 hr at 28°C. Finally the GFP positive colony numbers were recorded by manual counting to calculate the inhibition rate.

### Histology

All organs fixed in 10% formalin solution were embedded with paraffin and sectioned. Multiple sections defining the embedding direction and observation position in different organs were prepared for haematoxylin and eosin staining. The images were accessed by an Olympus BX51 system.

### Patients

The research ethics committee of Chinese PLA General Hospital (S2020-074-04) approved the study and all participants signed an informed consent according to the requirement of Ethics Commission. The studied subjects were from wuhan hankou hospital and huoshenshan hospital. All the patients were mild inpatients who were positive for nucleic acid test when enrolled. They met the inclusion requirements of the ethics committee. All subjects signed the informed consent. Since the patients had received systemic treatment in the local hospital at enrollment, for the sake of safety, on the premise that the original treatment plan was unchanged, the subjects were given extra LL-37 treatment by oral administration for 3 pills, 1 time/8 hours, and for 3 weeks. Baseline clinical information was collected, including demographic characteristic, clinical symptoms, blood test results, nucleic acid test results, lung CT results, etc., and follow-up was conducted once a week to evaluate the improvement of patients’ clinical symptoms and adverse reactions.

### Statistical Analysis

All data were shown by Mean ± SEM. Comparisons between groups were made with one-way ANOVA with Tukey’s post-hoc test or Student’s t-test. Repeated measurement analysis was made by both factors RM two-way ANOVA. p value < 0.05 was regarded as statistically significant.

## Results

### The recombinant pNZ8149-7×LL-37 construction and gene expression

As a food grade bacterial strains, lactococcus lactis was usually regarded as a very convenient living oral drug carrier which could transport protein drugs or vaccines. Here we designed a recombinant lactococcus lactis carrying LL-37(cas001), a human endogenous antiviral peptide to treat COVID-19 (Fig. 1A). In order to express the shorter target polypeptide, we combined signal peptide, LEISS enhancing sequence with seven tandem repeated sequences encoding mature human LL-37 under control of the nisin-inducible nisA promoter (Fig. 1B-C). The cas001 could secrete target protein (~34KD) into supernatant followed by addition of inducer Nisin(Fig. 1D). Therefore we successfully constructed LL-37 over-expressing lactococcus lactis.

### LL-37 impaired rVSV-SARS and rVSV-SARS-CoV2 infection in vitro

We investigated the inhibitory effect of LL-37 on virus entry into the target cells with the simulated viruses of SARS and SARS-CoV2 as follows. S protein is the main immunogen of coronavirus mediating virus infection. The rVSV-SARS and rVSV-SARS-CoV2 recombinant viruses were constructed by replacing the G protein of VSV with the S protein of SARS and SARS-CoV2, and the S protein was displayed on the surface of the VSV virus, respectively. The recombinant virus can simulate the infection process of SARS and SARS-CoV2.

To verify LL-37 effect in inhibiting SARS virus, packaged rVSV-SARS virus was diluted to 2000 ffu/ml as virus solution. LL-37 was then diluted to 0 ng/ml, 10 ng/ml, 100 ng/ml, 1000 ng/ml, 10000 ng/ml, 100000 ng/ml using virus solution to get different concentrations of treatment solution. After incubation for 30 minutes at the room temperature, different concentrations of treatment solution were used to treat vero cells in 96-well plates, with 100ul per well and 3 multiple wells for each concentration. The cells were placed in a 37°C cell incubator. The fresh medium containing 20 mM NH4Cl was added in place of treatment solution 2 hours after virus infection, and the cells were placed in cell incubator at 28°C. The number of cells with positive fluorescence signal was counted under fluorescence microscope 24 hours after infection. Finally, according to the comparison test results, the antiviral ability of LL-37 was identified. The maximum inhibition rate of LL-37 was 85% at the concentration of 10ug/ml (Fig. 2A). The ability of LL-37 to inhibit SARS-CoV2 infection was measured in similar manner. At a concentration of 100ng/ml, LL-37 inhibited the ability of SARS-CoV2 to invade Vero cells by 41.5% (Fig. 2B).

**Figure 2.**
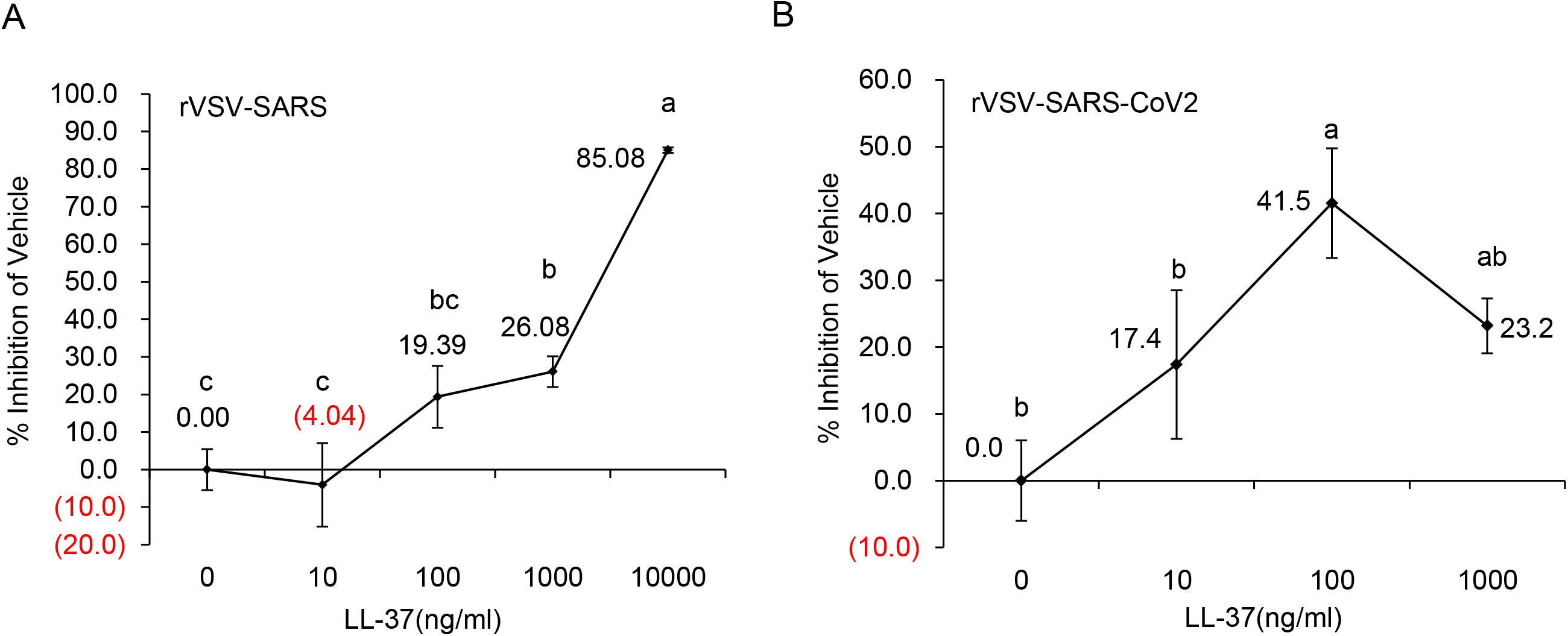
LL-37 impaired rVSV-SARS and rVSV-SARS-CoV2 infection in vitro. (A) Inhibition rate of LL-37 peptide on rVSV-SARS compared with vehicle(n=3). (B) Inhibition rate of LL-37 peptide on rVSV-SARS-CoV2 compared with vehicle(n=3). Multiple comparisons which were significant were represented with alphabetic notation, abc recorded as p value < 0.05.

### Results of the subchronic toxicity experiment showed that cas001 displayed good safety profiles

The acute and subchronic toxicity test of LL-37 was performed in rats to evaluate its safety. We produced special Lactococus Lactis with chloramphenicol resistance. Using culture plates with chloramphenicol resistance, we detected specially-made probiotic bacteria number in feces to determine its colonization in the gut. The chloramphenicol-resistant bacteria were administrated to mice by gavage (1 × 10^9^ CFU/mice), and the feces were collected at different time after the gavage (Fig. 3A). In order to determine the blood pharmacokinetic profile of LL-37, male SD rats (body weight 250-300g, 1 × 10^10^ CFU/animal, n=3) were given oral administration of cas001. The concentration of LL-37 in the blood before and after gavage was detected using ELISA kit (Hycult biotechnology Cat# HK321). Results showed that 2 hours after gavage, the concentration of LL-37 in the blood reached the peak (Fig. 3B).

**Figure 3.**
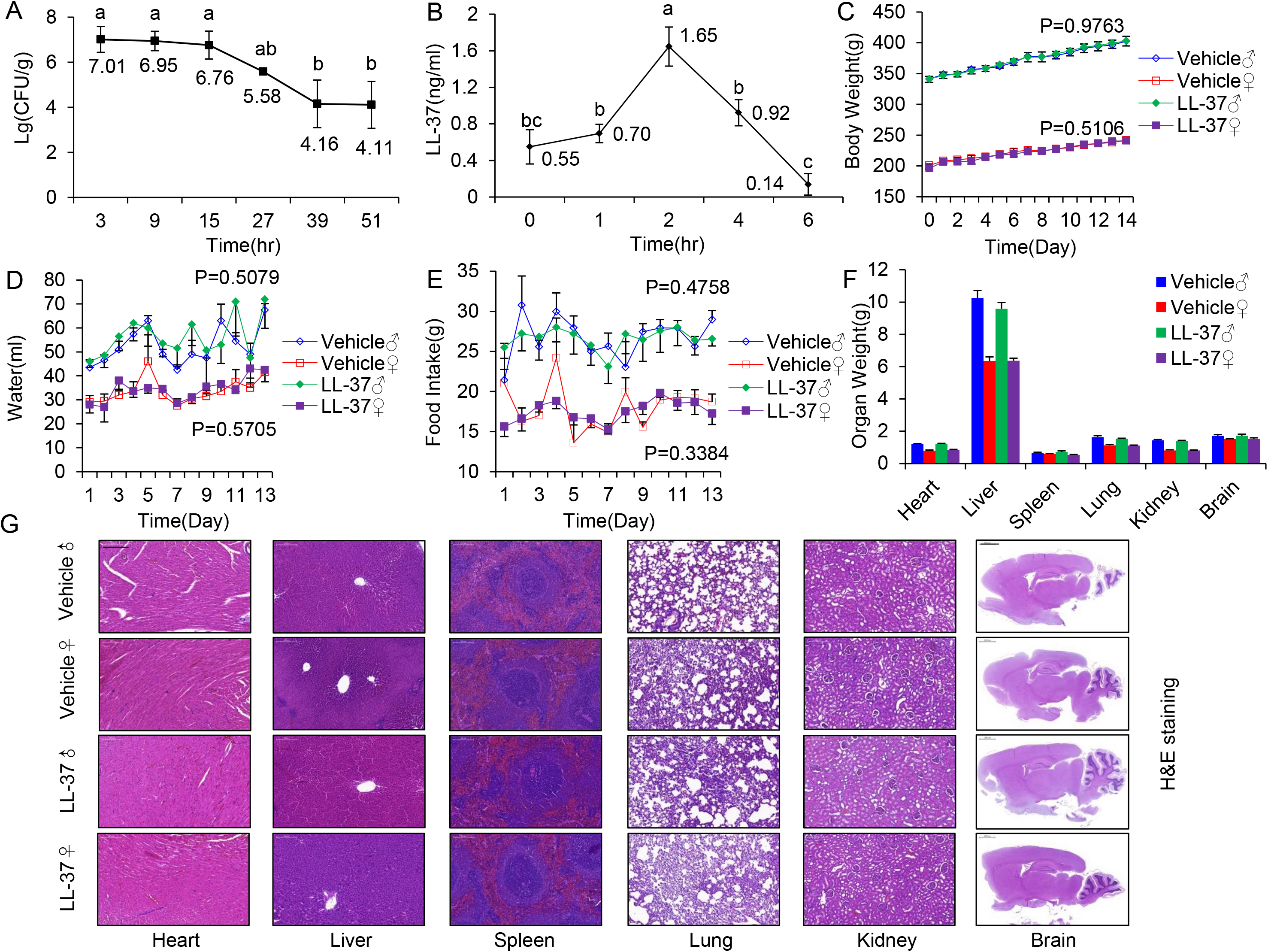
Oral administration of living biological drugs had little effect on the health of rats. (A) The bacterial counts of faeces at different time point after a single dose of oral administration of 1.0E9 frozen-dried powder with chloromycetin resistant probiotics. (B) The LL-37 concentration in rats’ venous blood after a single oral administration of 3.0E10 frozen-dried powder (n=3). (C) The body weight change, (D)water intake, (E) food intake, (F) organs weight (Vehicle v.s. ♂-37♂, n=5; Vehicle♀ v.s. LL-37♀, n=5) were monitored during subchronic toxicity experiment. (G) H&E staining of several organs sections from rats. Scale bars in images of heart, liver, spleen, lung and kidney refer to 200 μm and those in images of whole brain refer to 2000 μm.

In the animal safety evaluation experiment, 100 times of the expected clinical dose of drug was given to rats by oral gavage at the dose of 6 × 10^10^ CFU//kg/ day. The results showed that LD50 of LL-37 was more than 6 × 10^10^ CFU/kg/ day. Observation during 14 days of administration showed that all SD rats were generally in good condition both during and after administration and no abnormal performance was observed in SD rats (data not shown). The body weights showed no significant difference between the treatment group and the control group (Fig. 3C), indicating that the oral cas001 did not affect the weight gain. During the observation period, the food intake, water intake, organs weight and histological examination results of SD rats in treatment group were not significantly different from those of the control group (Fig. 3D-G), indicating that under this experimental condition, cas001 did not affect the appetite of experimental animals. The results of blood biochemistry examination indicated that there was no significant difference in liver, kidney function, myocardial enzyme concentration, blood lipid concentration and electrolyte concentration between the two groups of SD rats (all p values were > 0.05) (Table. 1).

**Table 1.** Serum biochemical detection in rats two weeks after gavage. Serum biochemistry detection of rats at the end of two weeks (Vehicle♂ v.s. LL-37♂, n=5; Vehicle♀ v.s. LL-37♀, n=5). ^*^represent p value<0.05.

| Group | TB<br>(μM) | DB<br>(μM) | TG<br>(mM) | TC<br>(mM) | AST<br>(U/L) | ALT<br>(U/L) | ALP<br>(U/L) | Cr (μM) | BUN<br>(mM) | LDH<br>(U/L) | CK (U/L) | Cl (mM) | K (mM) | Na (mM) | Ca (mM) | Mg<br>(mM) | P (mM) |
| --- | --- | --- | --- | --- | --- | --- | --- | --- | --- | --- | --- | --- | --- | --- | --- | --- | --- |
| Vehicle♂ | 0.96±0.12 | 0.78±0.09 | 0.37±0.09 | 1.07±0.08 | 126.74±6.83 | 59.56±3.28 | 160.50±5.01 | 22.70±0.68 | 4.00±0.32 | 440.40±50.26 | 561.62±34.74 | 102.24±0.84 | 6.02±0.18 | 145.04±0.61 | 2.53±0.03 | 1.13±0.03 | 2.86±0.14 |
| LL-37♂ | 1.3±0.1 | 1.18±0.07* | 0.29±0.03 | 1.37±0.08* | 127.38±10.55 | 47.20±3.83* | 152.62±22.32 | 22.38±0.48 | 4.65±0.33 | 571.86±71.69 | 661.02±93.22 | 101.24±0.29 | 5.68±0.15 | 145.06±0.36 | 2.54±0.01 | 1.04±0.04 | 2.81±0.07 |
| Vehicle♀ | 1.2±0.3 | 1.10±0.22 | 0.32±0.03 | 1.47±0.25 | 152.30±14.58 | 39.98±2.11 | 72.06±4.78 | 25.60±2.82 | 4.85±0.34 | 833.46±371.28 | 954.30±394.79 | 102.30±0.91 | 5.20±0.42 | 143.26±0.82 | 2.50±0.08 | 1.02±0.04 | 2.63±0.14 |
| LL-37♀ | 1.36±0.19 | 1.18±0.22 | 0.33±0.07 | 1.30±0.20 | 103.78±10.00* | 39.48±1.14 | 64.60±4.60 | 27.76±1.26 | 5.02±0.23 | 349.50±142.26 | 715.56±343.35 | 102.42±0.82 | 5.13±0.11 | 143.58±0.42 | 2.62±0.03 | 1.00±0.02 | 2.53±0.12 |

\*represent p value<0.05.

To assess pharmacokinetics in human individuals, four healthy adult volunteers were recruited to the study (Table. 2). During dose escalation study periods, the volunteers did not feel any discomfort while taking the cas001 capsules, such as fatigue, weakness, nausea, vomiting, etc. Two hours after oral administration, the concentration of LL-37 level was increased in healthy volunteers (#1volunteer who took highest dose of cas001, increased significantly, the others only increased slightly) (Table. 3). Accordingly, there was no significant change in liver and kidney function after oral cas001 medication (Table. 4). All these results taken together, cas001 displays an excellent safety profile in both preclinical trials and healthy volunteers assay.

**Table 2.**
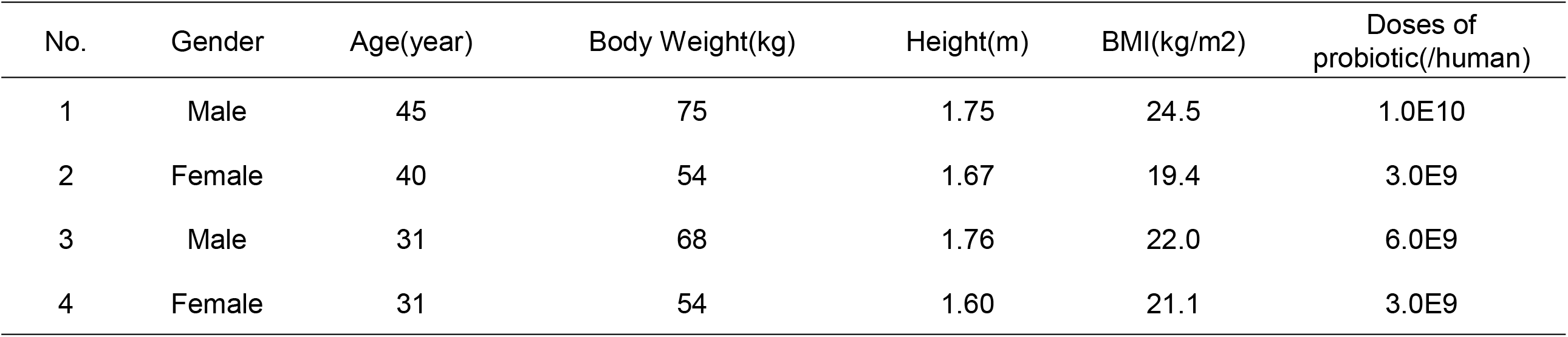
Demographic information of volunteers.

**Table 3.** The changes of serum LL-37 concentration before and after cas001 medication.

| LL-37 (ng/ml) | Time (0h) | Time (2h) |
| --- | --- | --- |
| Male1 | 54.1 | 65.7 |
| Female2 | 34.1 | 35.2 |
| Male2 | 39.4 | 42.3 |
| Female4 | 38.3 | 41.2 |

**Table 4.** Serum biochemical detection of volunteers.

| Treatment |  | Before |  |  |  | After 2 hr |  |  |  |
| --- | --- | --- | --- | --- | --- | --- | --- | --- | --- |
| Index | Standard values | Male1 | Female1 | Male2 | Female2 | Male1 | Female1 | Male2 | Female2 |
| ALT (U/L) | 0-40 | 12.6 | 7.8 | 22.4 | 6.6 | 10.2 | 7.1 | 25.1 | 3.8 |
| TP (g/L) | 55-80 | 77 | 77.6 | 72.3 | 73.6 | 76.2 | 76.7 | 73.2 | 74.1 |
| TB (μmol/L) | 0-21 | 9 | 3.4 | 5.3 | 3.3 | 9.1 | 4.1 | 6 | 3.4 |
| Tn (ng/ml) | 0-0.1 | <0.0030 | <0.0030 | 0.011 | <0.0030 | <0.0030 | <0.0030 | 0.011 | <0.0030 |
| γ-GT(U/L) | 0-50 | 12.1 | 10.9 | 25.1 | 10.8 | 12.7 | 10.3 | 26 | 11.8 |
| Cr (μmol/L) | 30-110 | 90.2 | 58.8 | 83.2 | 77.1 | 88.3 | 53.6 | 82.5 | 69.4 |
| Ca (mmol/L) | 2.09-2.54 | 2.41 | 2.47 | 2.4 | 2.42 | 2.44 | 2.54 | 2.43 | 2.47 |
| Mg (mmol/L) | 0.6-1.4 | 0.95 | 0.9 | 0.85 | 0.79 | 0.91 | 0.85 | 0.81 | 0.79 |
| Na (mmol/L) | 130-150 | 140.3 | 137.6 | 140.4 | 140.1 | 139 | 136.4 | 140.2 | 139.5 |
| CO2 (mmol/L) | 20.2-30.0 | 25.5 | 23.4 | 22 | 21.8 | 26.1 | 22.6 | 24.2 | 22.8 |
| LPS (U/L) | 13-60 | 27.1 | 44.4 | 53.1 | 26.4 | 28.4 | 40.3 | 49.1 | 26.6 |
| CK-MB (ng/ml) | 0-6.5 | 1.67 | 1.32 | 2.1 | 1.48 | 1.65 | 1.03 | 1.71 | 1.04 |
| AST (U/L) | 0-40 | 19 | 15.2 | 25.7 | 15.4 | 15.8 | 15.2 | 25 | 13.6 |
| ALB (g/L) | 35-50 | 51.2 | 49.3 | 46.8 | 49.6 | 52.3 | 47.3 | 48.3 | 48.8 |
| DB (μmol/L) | 0-8.6 | 4.2 | 2.2 | 2.5 | 2 | 4.2 | 2.3 | 2.8 | 2.2 |
| ALP (U/L) | 42-396 | 51 | 36.9 | 40.6 | 50.5 | 50.4 | 35.5 | 37.4 | 53.8 |
| BUN (mmol/L) | 1.8-7.5 | 4.79 | 6.64 | 5.45 | 5.79 | 4.55 | 6.23 | 5.73 | 6.06 |
| UA (μmol/L) | 104-444 | 316.2 | 251.7 | 377.8 | 370.9 | 313.5 | 249.6 | 372.4 | 364.5 |
| LDH (U/L) | 40-250 | 168.8 | 174.5 | 217.2 | 165.6 | 167.6 | 169.3 | 184.9 | 132.3 |
| P (mmol/L) | 0.89-1.6 | 1.37 | 1.43 | 1.14 | 1.51 | 1.35 | 1.51 | 1.26 | 1.25 |
| K (mmol/L) | 3.5-5.5 | 3.99 | 4.29 | 3.89 | 4.42 | 3.95 | 4.24 | 4.01 | 4.2 |
| Cl (mmol/L) | 94-110 | 100.4 | 101 | 104.8 | 102.8 | 100.2 | 101.5 | 103.1 | 103.5 |
| AMS (U/L) | 0-150 | 71 | 59.8 | 69.9 | 65 | 69.7 | 59.3 | 68.4 | 64.7 |
| Mb (ng/ml) | 0-75 | <21.0 | <21.0 | 76.7 | <21.0 | <21.0 | <21.0 | 41.7 | <21.0 |
| LDH-MB (U/L) | 17-72 | 40 | 38.5 | 48.3 | 42.8 | 38.5 | 36.5 | 38.3 | 32.3 |

### Cas001 significantly improved symptoms of COVID-19 patients

With the approval of ethics committee of Chinese PLA General Hospital (Ethics no.: s2020-074-04), the investigator-initiated clinical trial (ITT) was carried out, which mainly focused on safety assessment and small-scale, single-arm, exploratory safety study was performed. A total of 11 eligible patients with mild symptoms were enrolled in Wuhan Hankou hospital and Huoshenshan hospital. They were enrolled voluntarily and all patients signed informed consent. Among them, there were 5 males and 6 females, aged 55 ± 12 (36-70) years old, and the duration from onset to medication enrollment was 35±19 (5-68) days. 6 patients were nucleic acid positive and 5 patients were nucleic acid negative when they were enrolled. All patients received the cas001 treatment according to requirement (3 capsules/time, three times a day for 3weeks), with an average follow-up time of 33±15 days (Table. 5).

**Table 5.** Baseline patient characteristics.

| Name | Gender | Age | Time from onset to enrollment (d) | Clinical classification | Nucleic acid test at enrollment |
| --- | --- | --- | --- | --- | --- |
| Patient #1 | F | 57 | 5 | mild | + |
| Patient #2 | F | 70 | 30 | mild | + |
| Patient #3 | M | 70 | 30 | mild | + |
| Patient #4 | M | 51 | 37 | mild | + |
| Patient #5 | M | 57 | 35 | mild | + |
| Patient #6 | M | 36 | 68 | mild | - |
| Patient #7 | F | 42 | 5 | mild | - |
| Patient #8 | F | 65 | 36 | mild | + |
| Patient #9 | M | 65 | 33 | mild | - |
| Patient #10 | F | 40 | 47 | mild | - |
| Patient #11 | F | 57 | 58 | mild | - |

At the time of enrollment, the patients’ symptoms included three categories, among which the positive systemic symptoms mainly included fever, fatigue and myalgia. After treatment, the improvement rate of systemic symptoms was 100%, and the time from medication to the complete remission ranged from 5 to 33 days; Respiratory symptoms included pharyngalgia, cough, chest distress, shortness of breath, which were improved completely 7-14 days after treatment with improvement rate up to 100%; Gastrointestinal symptoms including abdominal discomfort, nausea, poor appetite, and diarrhea were completely improved within 2-18 days after treatment. Among all symptoms, gastrointestinal symptoms were improved fastest with the shortest improvement time of 2 days (Table. 6).

**Table 6.** Outcomes in the intention-to-treat population.

|  | Systemic symptoms |  |  | Respiratory symptoms |  |  |  |  | Gastrointestinal symptoms |  |  |  |
| --- | --- | --- | --- | --- | --- | --- | --- | --- | --- | --- | --- | --- |
| Observation index | fever | weak | myalgia | pharyngalgia | cough | chest distress | dyspnea | shortness of breath | abdominal discomfort | nausea | poor appetite | diarrhea |
| Positive cases | 2 | 6 | 4 | 2 | 5 | 3 | 1 | 3 | 7 | 2 | 3 | 4 |
| Average duration of improvement (day) | 4 | 14 | 16 | 6 | 17 | 12 | 7 | 9 | 10 | 5 | 14 | 12 |

Among the 11 patients enrolled, 5 were positive for nucleic acid test before medication, 6 were negative. After treatment, 4 of the 5 positive patients were negative for nucleic acid test. The duration from taking the medicine to recovery was 19, 17, 9 and 17 days respectively. All patients received lung CT examination before the enrollment and nine of them showed abnormal lung CT results. During the follow-up period, a total of 4 cases completed the reexamination of lung CT, 3 cases of them have been completely improved, the duration from medication to complete improvement was 44, 48, 50 days, respectively; Another 1 case improved 75%, costing 17 days from medication to improvement. The remaining 5 patients did not complete the pulmonary CT reexamination due to home isolation/quarantine place isolation after discharge (Table. 7).

**Table 7.** Nucleic acid detection and lung CT examination.

| Observation index | Positive cases at enrollment | Reexaminaion cases * | Completely recovered cases | 75% recovered cases (CT) | Average medication time(d) | Negative conversion rate/complete absorption rate(%) | Partial absorption rate(%) |
| --- | --- | --- | --- | --- | --- | --- | --- |
| Nucleic acid detection | 5 | 5 | 5 | - | 15 | 100 | - |
| Lung CT | 9 | 4 | 3 | 1 | 47 | 100 | 100 |

\* The remaining 5 patients did not complete the pulmonary CT reexamination due to home isolation/quarantine place isolation after discharge

11 patients enrolled showed good compliance, tolerance, subjective feeling and actively interacted with the doctors. None of the patients had any adverse reactions, and the observation of adverse reactions was performed with reference to literatures^21^. Living bacteria drugs may cause four types of adverse reactions theoretically, including gastrointestinal related adverse reactions, infection, harmful metabolic effects, excessive immune stimulation of susceptible individuals and gene transfer^22^. There was no clinical evidence for the occurrence of the latter two, and the main manifestations of the other three types of adverse reactions including infection^23-29^, harmful metabolic effect^30^, gastrointestinal related adverse reactions^22^ were as follows (Table. 8). During the observation of this clinical study, none of the patients presented adverse reactions shown in table 4 or any other ones.

**Table 8.** Summary of probiotic-related adverse reactions.

| Category | Detailed adverse reactions | Cases observed |
| --- | --- | --- |
| Infection 【Ref 23-29】 | bacteremia, fungemia, sepsis,<br>endocarditis | 0 |
| Harmful metabolic effects 【Ref 30】 | intestinal ischemia, D-lactic acid<br>poisoning | 0 |
| Gastrointestinal related adverse<br>reactions 【Ref 22】 | diarrhea, abdominal distension,<br>abdominal discomfort, stool shape<br>change, nausea, vomiting | 0 |

## Discussion

There is a report indicating that although gastrointestinal symptoms were uncommon, still a part of patients showed gastrointestinal symptoms, such as diarrhea (3.8%) and vomiting (5.0%)^31^. In line with this observation, there are numbers of studies demonstrating that SARS-CoV-2 can be detected in the gastrointestinal tract^32, 33^ Since the SARS-CoV-2 spike glycoprotein binding to ACE2 receptor is a critical step for virus entry, investigation of the expression pattern of ACE2 in host specific target cells or organs is particular important. Accordingly, a report demonstrated that ACE2 is rarely expressed in esophageal epithelium but is abundantly distributed in the cilia of the glandular epithelia^33^. These results imply that we cannot exclude the possibility of fecal-oral transmission.^34^

Lactococcus lactis could be used to treat GIT related diseases such as colitis and inflammatory bowel disease (IBD) due to its survival ability in gastrointestinal tract (GIT) and its immunomodulatory properties. In fact, engineered Lactococcus lactis secreting interleukin-10 (IL-10) was used to treat murine colitis^19^. Although clinical outcome was not as promising as hoped in human crohn’s disease^20^, similar clinical trials addressed concerns related to release of lactococcus lactis. It should be noted that we should consider the properties of lactococcus lactis in the mucosal immunity, besides systemic target protein secretion by GMOs. Interestingly, a report demonstrated that lactococcus lactis induced mucosal IgA which can form a barrier to pathogens on the mucosal surface^35^. Whether cas001 could also induce IgA mucosal immunity needs further investigation.

According to the scope of study and number of cases approved by the medical ethics committee, the first phase of the single-arm, symptomatically oriented investigator initiated clinical trials (ITT) which enrolled 11 patients have been successfully completed. From the results of research and follow-up, patients were well tolerated, characterized by good compliance and feeling after taking cas001. The adverse reactions reported in the symptomatology and probiotics related literatures include: gastrointestinal related adverse reactions, infection, and harmful metabolic effects, while all of the above were not observed in the study. Therefore, from this small scale exploratory research, the good safety of cas001 was preliminarily verified.

All patients enrolled are mild in symptoms and in convalescence, the ratio of male: female is appropriate, the duration from the onset to the enrollment is at the average of 34 days. After the medication, systemic symptoms, respiratory symptoms, gastrointestinal symptoms were improved to varying degrees, among them gastrointestinal symptoms were improved significantly. The improvement rate of respiratory symptoms was 100%, all patients showed improvement during the follow-up period, and no recurrence of the above symptoms occurred during the follow-up period after drug withdrawal. There were 5 patients with positive nucleic acid test results at the beginning of medication, and 4 patients turned negative after an average of 15 days medication. Another patient was in home isolation and did not receive reexamination. Whether the change of the nucleic acid test results is related to cas001 is not easy to infer at present. In terms of the improvement of lung CT results, 3 cases got completely improved and 1 case got 75% improved. It took 7days for recovery from mild inflammation to complete absorption of inflammation. Whether the cas001 improves or accelerates healing process remains to be investigated. Overall, cas001 shows definite effect in the improvement of gastrointestinal symptoms and is possible to have effects in improving the systemic symptoms and respiratory symptoms and may play a role in the improvement of results of nucleic acid test and lung CT test.

This study is limited to a small scope and mainly focuses on the safety exploration. The epidemic of novel coronavirus in China is in the clearance phase, so the number of medical records included from the above two centers is very limited. What’s more, there are few clinical cases related to cas001, which is a major defect of current clinical study. In addition, all patients were discharged from the hospital 3-5 days after enrollment, and were isolated at home or in a quarantine place, resulting in incomplete blood and CT follow-up data. The follow-up time is relatively short, and further evaluation is needed to see whether there are new safety problems in the middle phase (3-6 months) and the adverse reactions in middle and later phase. While, since Lactococcus lactis is not colonized in the body and has a short metabolic cycle and did not cause any adverse effects in short term observation, there is little possibility that cas001 would cause adverse reactions in middle and later phase.

This small scale exploratory study suggested that the oral cas001 has good clinical tolerance and significant safety. Cas001 has positive effect in the improvement of symptoms and nucleic acid clearance in patients with mild symptoms, especially in the improvement of gastrointestinal symptoms, suggesting that this drug could be used to treat patients with mild symptoms as an auxiliary drug or as a single cure for clinical application. Its efficacy and safety still need to be further confirmed by prospective, large sample, multi-center, randomized, double-blind, controlled studies and long-term follow-up.

### Limitations

Current study has several following limitations. Firstly, primary purpose of current clinical trial is assessment of safety profiles. In this regard, we did not performed dose escalation study, therefore, optimal clinical dose of cas001 is not yet fully determined. Secondly, this was a small scale single arm clinical trial without control. It is hard to reach conclusion of clinical outcomes related to the cas001, although changes of several symptoms indicating encouraging findings. Thirdly, before taking cas001, these self-quarantined patients with mild symptoms had received other antiviral drugs. It is unclear if these patients would be improved without cas001 treatment. Fourthly, it is hard to measure circulating LL-37 in patients due to viral contamination. Finally, we also could not directly quantify viral road because of limited number of patients available in the study and most of them were with mild symptoms and nucleic acid positive and were self-quarantined.

## Conclusions

In current preliminary uncontrolled study enrolling 11 patients with COVID-19, administration of cas001 has promising clinical outcomes. The small sample size and uncontrolled study design preclude a definitive statement about the potential effectiveness of cas001, and further evaluation in large-scale clinical trials is required.

## Data Availability

primer sequences are available from corresponding author upon request.

## Acknowledgement

This work was supported by the National Key Research and Development Program of China (2017YFC1001001), the National Natural Science Foundation of China (81770834), the Youth Program of National Natural Science Foundation of China (31900830) from the Chinese Academy of Sciences and the China Postdoctoral Science Foundation Funded Project (2018M640182). We would also like to thank Professor Cheng Wang from Harvard University for his great support for this study.

## Author contribution

H. Zhang, X. Jiang, C. Yan, J. Jiao, C. Zhao and H. Li together carried out the experiments. H. Zhang, Y. Zhao, X. Jiang, Li Chen and M. Dong together wrote the manuscript. Y. Zhao, Z. Luan, W. Chen, C. Feng, L. Tian, E. Qin, J. Mu, C. Li collected the clinical data and made revisit. T. Zeng, S. Feng, S. Wang, X. Guan, T. Li and H. Yu contacted hospitals and patients and analyzed the results. A. Zheng, W. Jin, G. Sun conceived the project. All authors were involved in editing the paper and had final approval of the submitted and published versions.

